# Whole-Body Muscle MRI Characteristics of LAMA2 Gene Mutation Congenital Muscular Dystrophy Children

**DOI:** 10.1101/2021.03.27.21254399

**Authors:** Hossam M. Sakr, Nagia Fahmy, Nermine S. Elsayed, Hala Abdulhady, Tamer A. El-Sobky, Amr M. Saadawy, Christophe Beroud, Bjarne Udd

## Abstract

Merosin-deficient or LAMA2-related congenital muscular dystrophy (CMD) belongs to a group of muscle diseases with an overlapping diagnostic spectrum. MRI plays an important role in the diagnosis and disease-tracking of muscle diseases. Whole-body MRI is ideal for describing patterns of muscle involvement. Our purpose is to analyze the pattern of muscle involvement in merosin-deficient CMD children employing whole-body muscle MRI. Ten children with merosin-deficient CMD underwent whole-body muscle MRI. We used a control group of other hereditary muscle diseases, which included 13 children. Overall, 37 muscles were graded for fatty infiltration using Mercuri scale modified by Fischer et al 2008. The results showed a fairly consistent pattern of muscle fatty infiltration in index group, which differed from that in control group. There was a highly statistically significant difference between the two groups in regard to the fatty infiltration of the neck, serratus anterior, rotator cuff, deltoid, forearm, gluteus maximus, gluteus medius, gastrocnemius and soleus muscles. Additionally, results showed relative sparing of the brachialis, biceps brachii, gracilis, sartorius, semitendinosus and extensor muscles of the ankle in index group. There is evidence to suggest that whole-body muscle MRI can become a useful contributor to the differential diagnosis of merosin deficient CMD.

**Study Highlights:**

1. Whole-body muscle MRI is a useful contributor to the differential diagnosis of merosin-deficient (LAMA2-related) congenital muscular dystrophy.
2. The study demonstrated the presence of a fairly characteristic pattern of whole-body muscle MRI in children with merosin-deficient CMD in terms of sensitivity and specificity.
3. The study may be considered as a preliminary step of arriving at an MRI classification system for children with merosin-deficient CMD.

## Introduction

Congenital muscular dystrophy (CMD) is a group of clinically and genetically heterogeneous muscle disorders that usually presented early in life with muscle weakness, hypotonia and in which muscle biopsy reveals dystrophic myopathy [1-3]. Based upon specific clinical features, CMD can be classified into three main subcategories namely (a) collagenopathies or collagen VI-related myopathies, (b) merosinopathies or merosin-deficient CMD also known as Laminin alpha2 gene (LAMA2)–related CMD or CMD1A and (c) dystroglycanopathies [1]. Accurate and timely diagnosis of different subtypes of CMDs is important to prognosis, genetic counseling and institution of appropriate treatment and it includes many aspects as molecular diagnosis, pre-natal counseling, prognosis, the availability of clinical trials, and treatment, which in turn require multi-disciplinary cooperation [1, 2]. Merosin-deficient CMD or (LAMA2) –related CMD is characterized by severe skeletal muscle involvement resulting in hypotonia, inability to walk, joint contractures and normal or near normal cognitive brain functions [2, 3]. In addition to the main clinical features, the presence of white matter abnormalities on brain MRI and total or partial absence of merosin on immunohistochemistry of muscle biopsy with a high serum creatine kinase argues for merosin-deficient CMD [1, 2].

Generally speaking, dystrophic muscle changes in muscle dystrophies and primary myopathies have a specific clinical and muscle magnetic resonance imaging (MRI) pattern highly suggestive for the underlying protein defect and the responsible gene [1, 3, 4]. Therefore, muscle MRI and specifically whole-body muscle MRI is currently regarded as a valuable non-invasive diagnostic tool to reveal distinct pathologic patterns of a wide array of genetic muscle diseases including congenital muscular dystrophies [5, 6], congenital myopathies [4, 7], dystrophinopathies as Duchenne muscular dystrophy [8-10], limb-girdle muscular dystrophies as of facioscapulohumeral muscular dystrophy [11-14], among others.

MRI has many advantages for muscle imaging, as it is not operator dependent, can image superficial and deep structures with equal efficacy and it is possible to image muscles at multiple and large areas of the body, thus it is ideal for describing patterns of muscle involvement [15]. MRI also helps in the selection of the muscle to be biopsied in cases of absent clear muscle weakness and in advanced cases when muscle tissue is replaced by fat, lacking radiation, MRI is ideal in disease progression follow up [16]. Each MRI examination is formed of multiple sequences, some allow detection of fatty infiltration (T1 and 3-point Dixon sequences) and others detect edema (T2 and STIR sequences) [16].

Whole-body muscle MRI protocols were introduced based on advances in MRI acquisition making evaluation of the pattern of involvement of the entire body muscles possible as well as evaluation of organs other than the skeletal muscles e.g. the heart muscle and brain [14]. MRI can quantify the degree of muscle fatty infiltration using semi-quantitative scales such as the Mercuri scale modified by Fischer et al in 2008 [17]. Additionally, muscle diffusion tensor imaging has been shown to be potentially useful in early detection of pathological changes in muscles in Pompe disease that otherwise appear healthy on conventional MRI [18].

The extreme paucity of studies –case reports in isolation or amongst a heterogeneous series-that report the muscle MRI characteristics in merosin-deficient CMD [19-22] inspired us to perform this study. In this study we present the first literature report on a homogenous sample of merosin-deficient CMD children. The objective of which is to identify the pattern and extent of muscle involvement in a subset of children with clinically and genetically confirmed merosin-deficient CMD.

## Material and Methods

In this retrospective study, ten children with merosin-deficient CMD (mean age is 10.3 SD (+/-) 2.8 years, 3 boys and 7 girls) received a detailed neuro-orthopedic examination, muscle biopsy with immunohistochemistry, and serum CPK to confirm the diagnosis. Eight children received a molecular study of the LAMA2 gene, which ascertained the diagnosis of merosin-deficient CMD. Children received whole-body muscle MRI to detail the pattern of muscle involvement in all extremities and the trunk. Consequently, the final diagnosis was verified by establishing a correlation between the neuro-orthopedic manifestations, muscle immunohistochemistry examination, brain MRI and molecular/genetic analysis.

Whole-body muscle MRI examinations were performed using a Philips Achieva 1.5 Tesla scanner (Philips, Netherlands). We performed T1W & STIR axial images for the whole-body (from head down to the leg) using the following parameters; T1WI: repetition time: 961ms, echo-time: 17 ms and thickness 5mm, STIR images: repetition time 5300 ms, echo time 64 ms, inversion time 165 ms in 5 mm slices. Each MRI examination was done after at least 30 minutes of patient rest to avoid the effects of activity, the total acquisition time was approximately 30 minutes. Fatty infiltration of muscle was graded on axial T1-weighted images using Mercuri scale modified by Fischer et al 2008 [17]. Image interpretation was performed separately by two experienced musculoskeletal radiologists. Their results were discussed with the rest of the authors and average was taken finally.

We used a control group of other hereditary muscle diseases which included 13 children (mean age was 13 SD (+/-) 5.5 years, 8 boys and56 girls); three with titin deficiency, two with congenital myopathy, two with fascio-scapulao-humeral muscular dystrophy, two with sarcoglyconopathy, two with limb girdle muscular dystrophy, non-specified, one with collagen VI-related myopathy and one with merosine-positive congenital muscular dystrophy.

### Statistical methods

Data was revised for its completeness and consistency. Data entry was done on Microsoft Excel workbook. Quantitative data was summarized by Mean, standard deviation while qualitative data was summarized by frequencies and percentages. The program used for data analysis is IBM SPSS statistics for windows version 23 (IBM Corp., Armonk, NY, USA). Chi-square test, student t test, and Pearson correlation coefficient were used in analysis of this study. Kappa test was done to measure level of agreement. A “P value” of less than 0.05 was considered statistically significant.

## Results

We selected 37 muscles all over the body for analysis and classified into 12 groups [Table 1]. The index group showed a fairly constant pattern of muscle involvement, all of them showed bilateral nearly symmetrical pattern [Figure 1] with noticeable pattern of muscle involvement and sparing e.g. the shoulder region showed affection of the rotator cuff muscles with relative sparing of the deltoid muscle [Figure 2]. The trunk showed specific predilection to involve the serratus anterior, erector spinae and inter costal muscles with relative sparing of the pectoralis major muscle. There was also a specific predilection to involve the gluteal region especially the gluteus medius and minimus muscles [Figure 3], the anterior and medial thigh muscles especially adductor brevis with relative sparing of the sartorius and gracilis, the hamstring muscles with relative sparing of the semitendinosus [Figure 4]. The leg region showed affection of all of the calf muscles in addition to the peroneal muscles of the lateral compartment [Figure 5]. The results are presented in [Table 2].

**Table 1.**
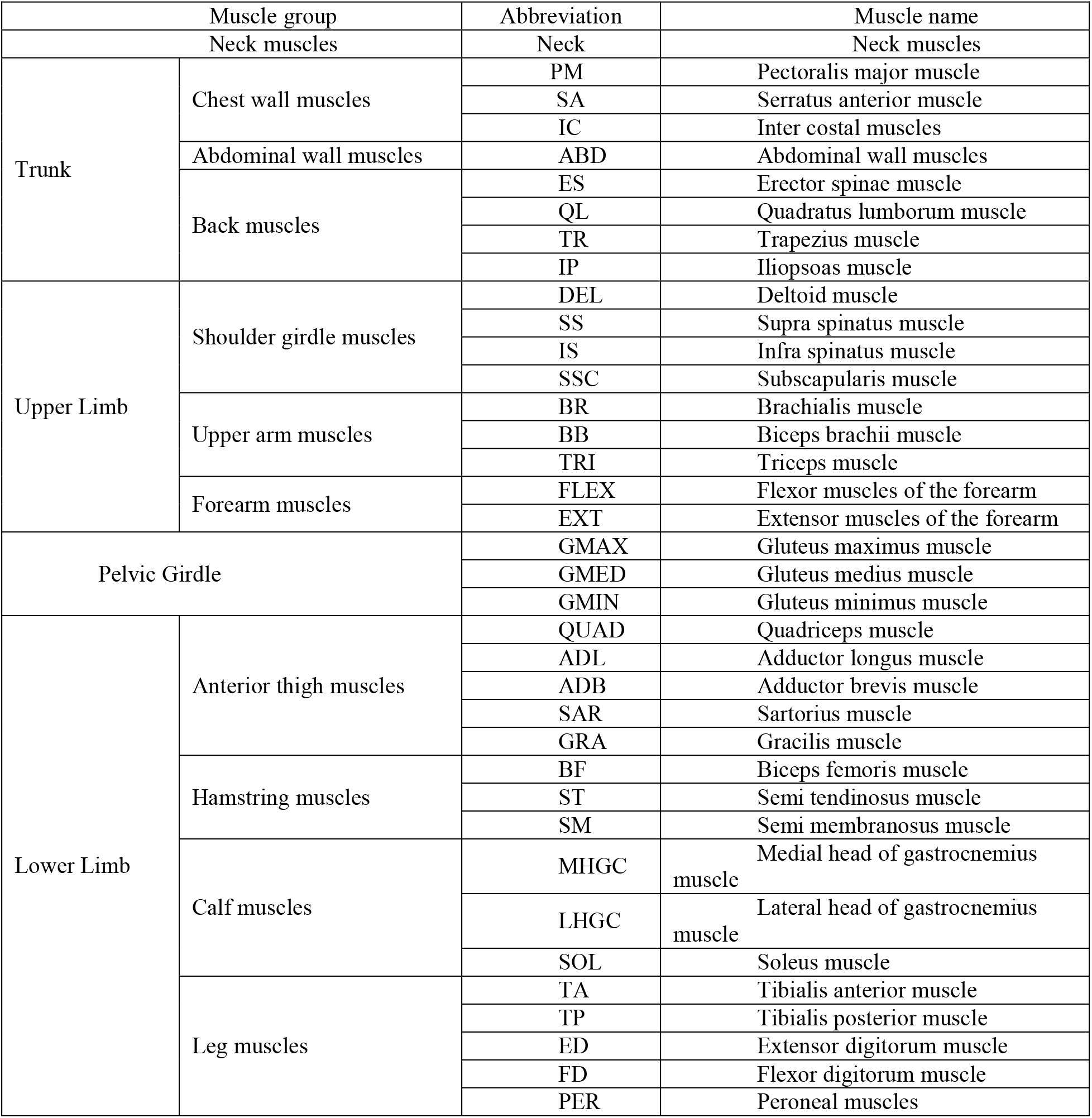
Muscles involved in the study, their classifications, and their abbreviations

**Table 2.**
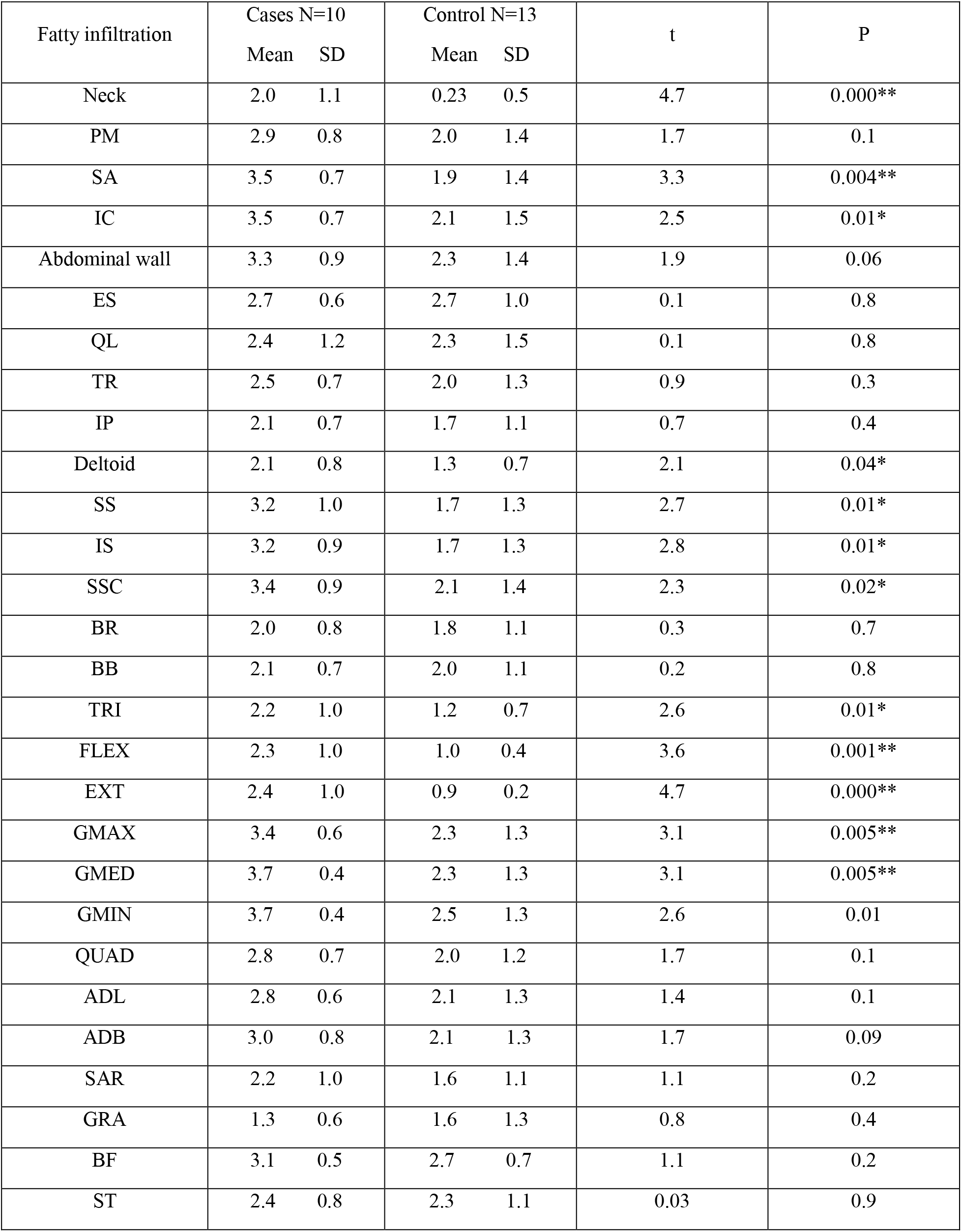
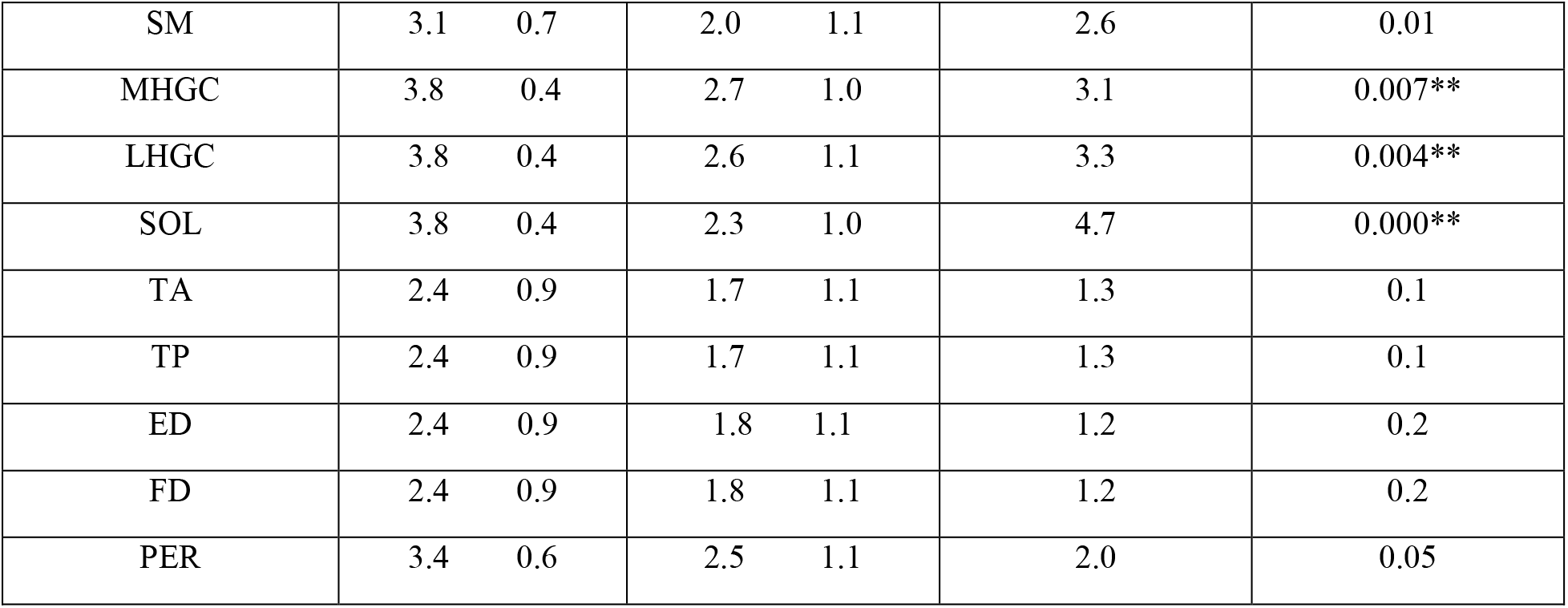
Mean score of fatty infiltration in the examined muscles (37 muscle) in both index and control groups with t and p values. * P<0.05 significant; **P<0.01 highly significant.

**Figure 1.**
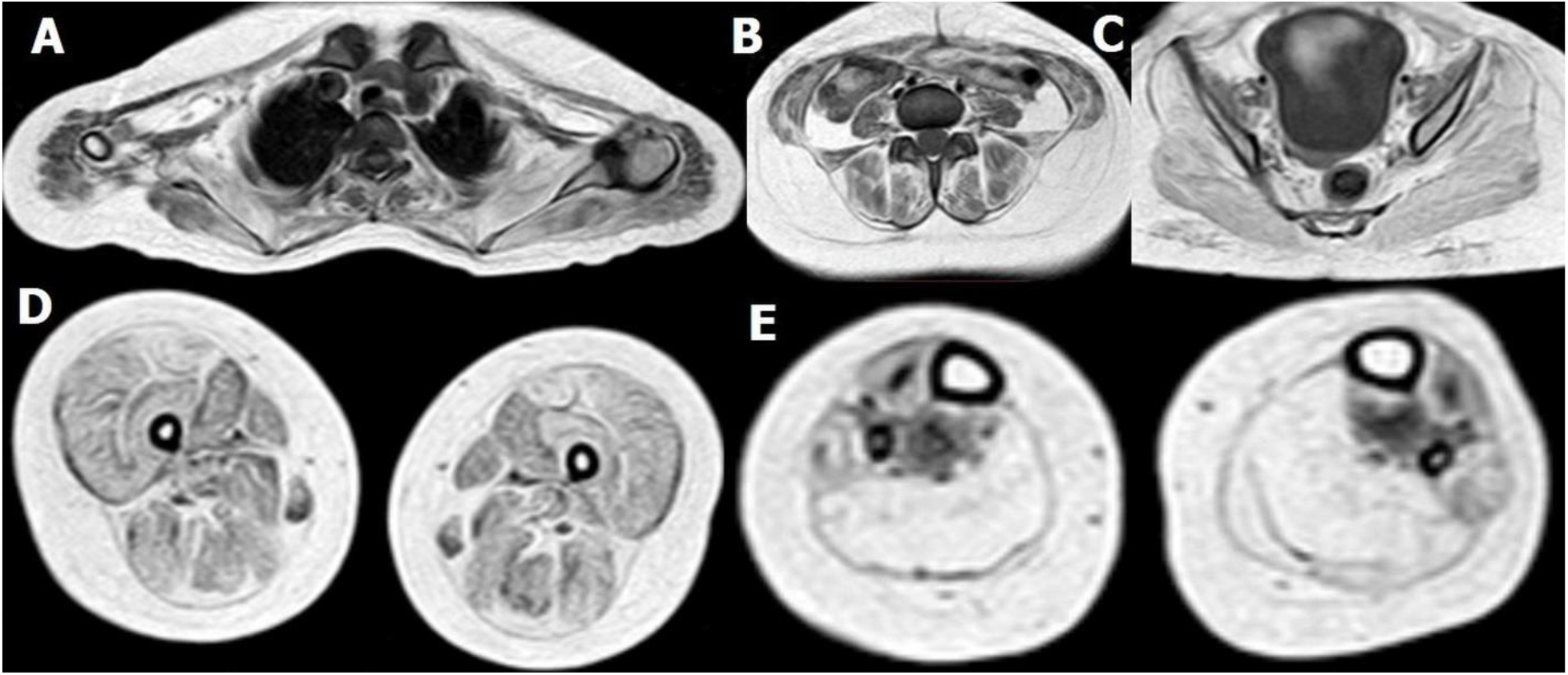
A patient with LAMA 2 mutation CMD: Selected axial T1WI of different regions showing bilateral nearly symmetrical pattern of muscle affection and fatty degeneration in shoulder A (mainly subscapularis), para spinal B, gluteal C (mainly gluteus maximus), thigh D (mainly quadriceps) & calf E (mainly posterior compartment muscles) regions.

**Figure 2.**
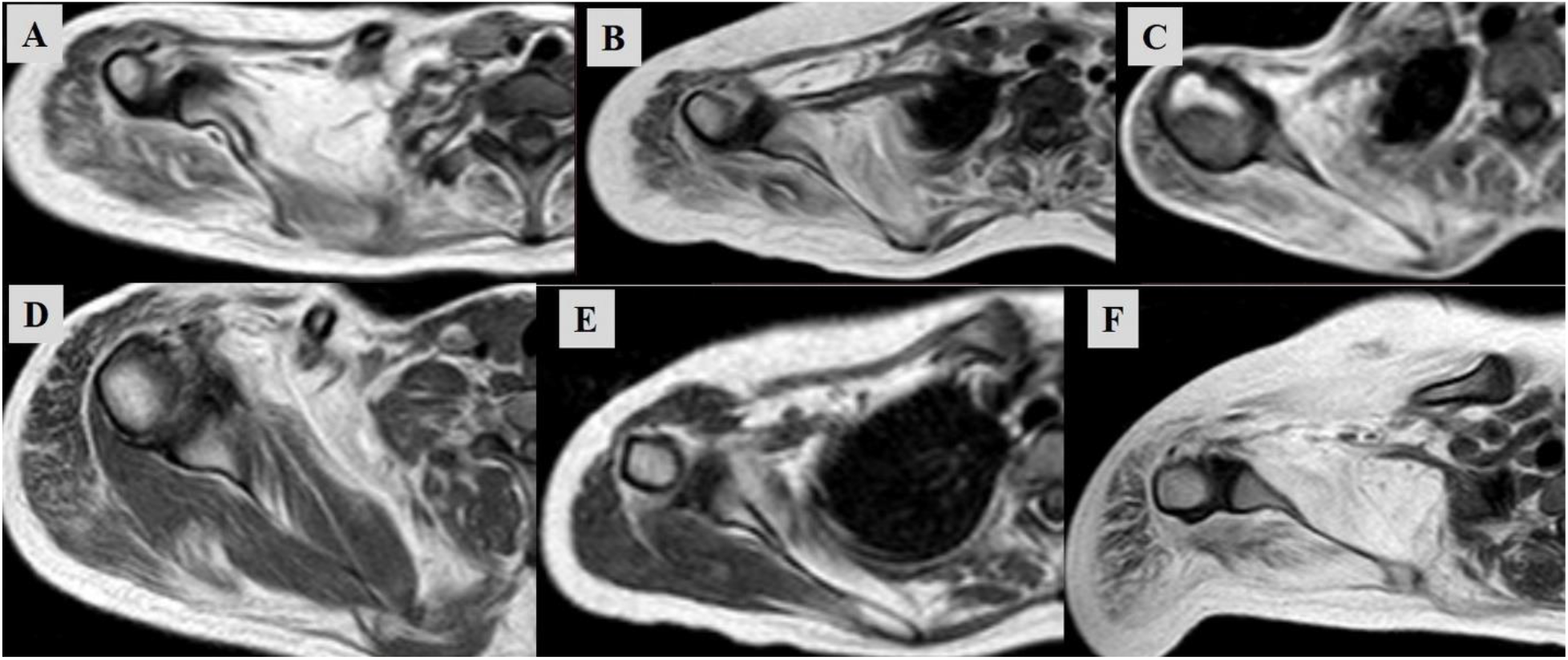
Selected Axial T1WI of the shoulder region in three different patients with LAMA 2 mutation CMD (A, B, C) & three different control patients (D: collagen 6 deficiency, E: titin deficiency, F: sarcoglyconopathy) showing affection of subscapularis and to a lesser degree infra-spinatus muscle with relative sparing of the deltoid muscles is evident in the three patients in contrary to the control cases.

**Figure 3.**
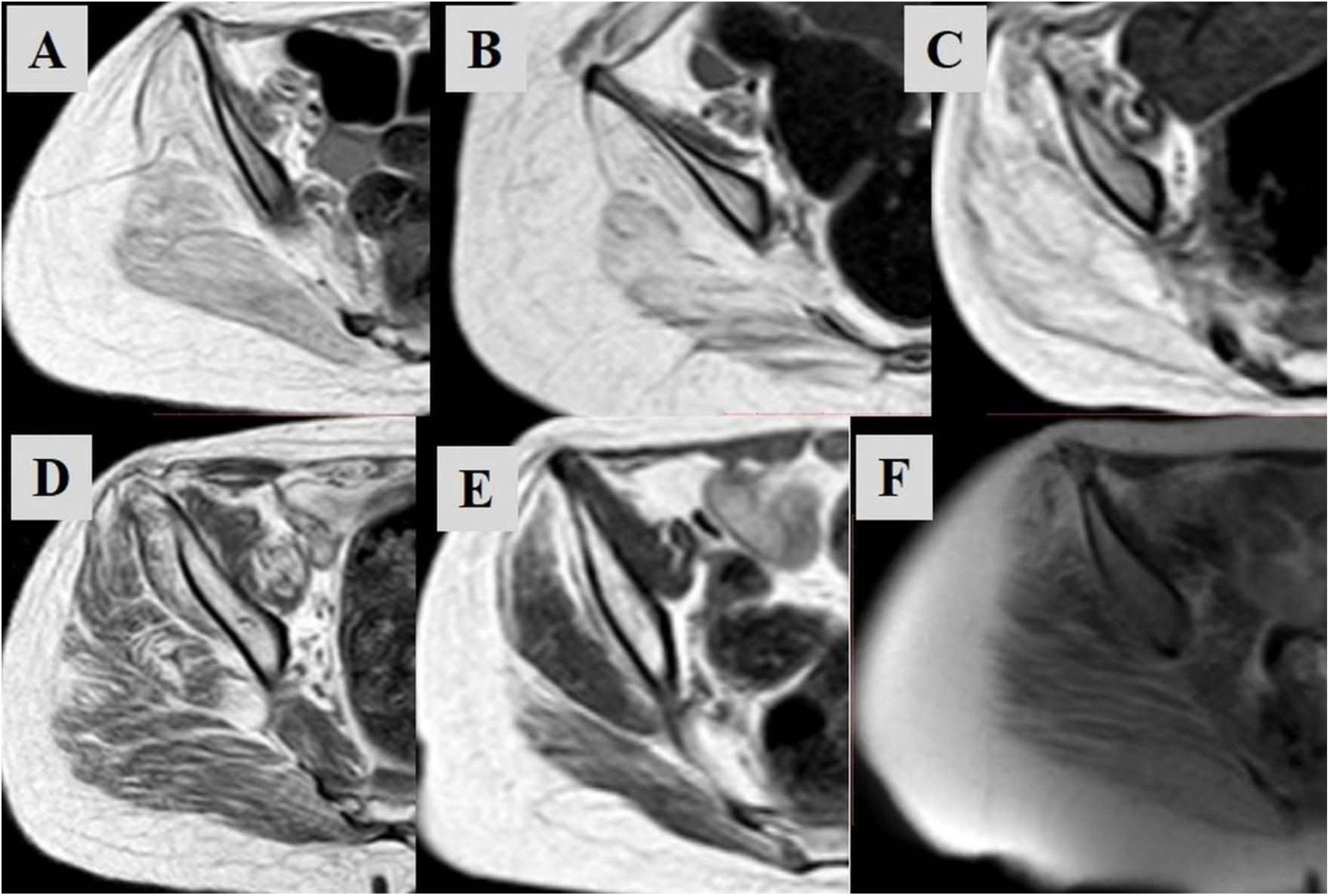
selected axial T1WI of the gluteal region in three different patients with LAMA 2 mutation CMD (A, B, C) & three different control (D: collagen 6 deficiency, E: titin deficiency, F: sarcoglyconopathy) showing moderate to severe grades of affection of the glutei muscles are evident in the three patients in contrary to the control cases.

**Figure 4.**
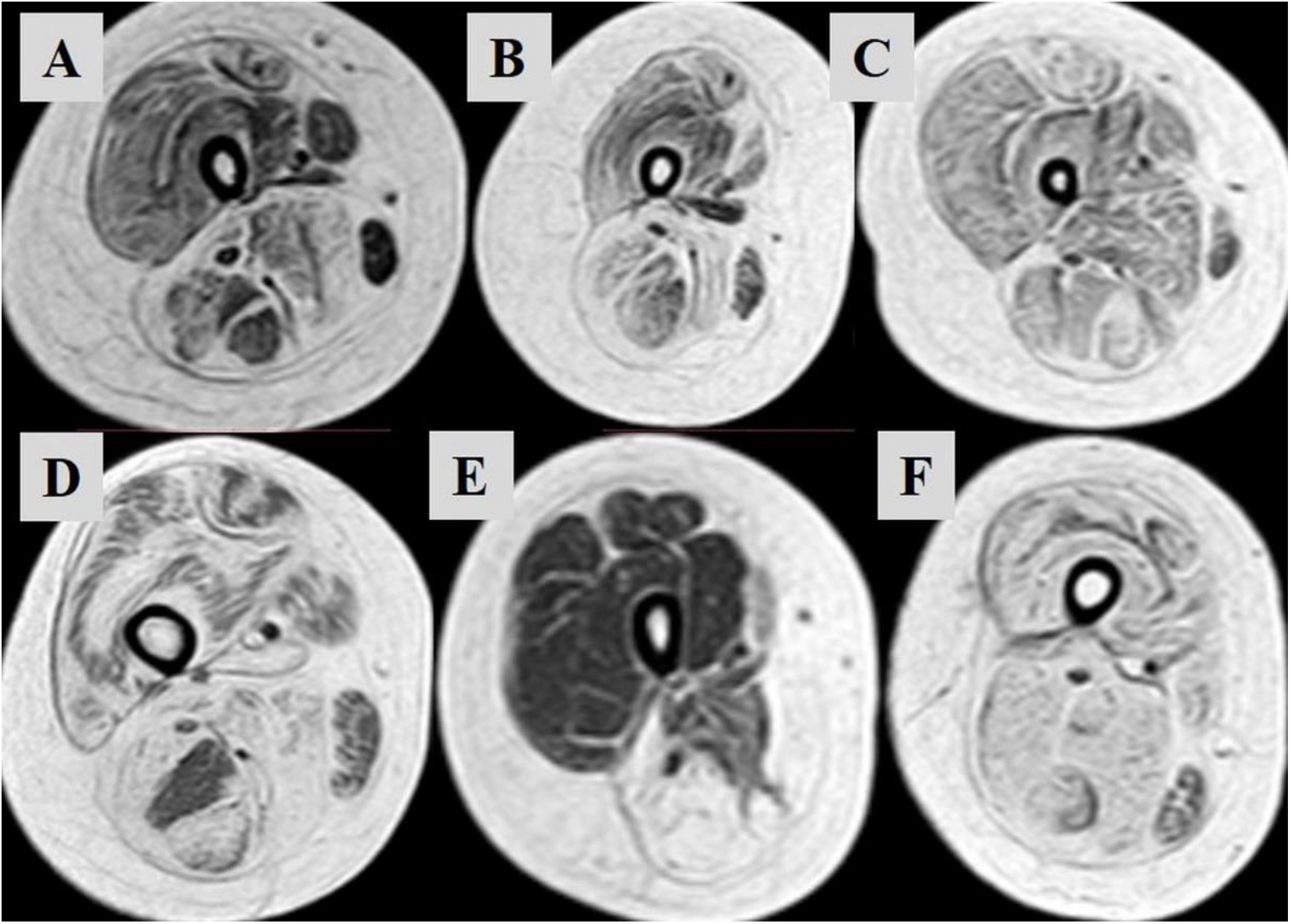
Selected Axial T1WI of the thigh in three different patients with LAMA 2 mutation CMD (A, B, C) & three different control (D: collagen VI deficiency, E: titin deficiency, F: sarcoglyconopathy) showing affection of the hamstring muscles with relatively spared gracilis, sartorius and semi tendinosus muscles, a pattern that differs from that in the control.

**Figure 5.**
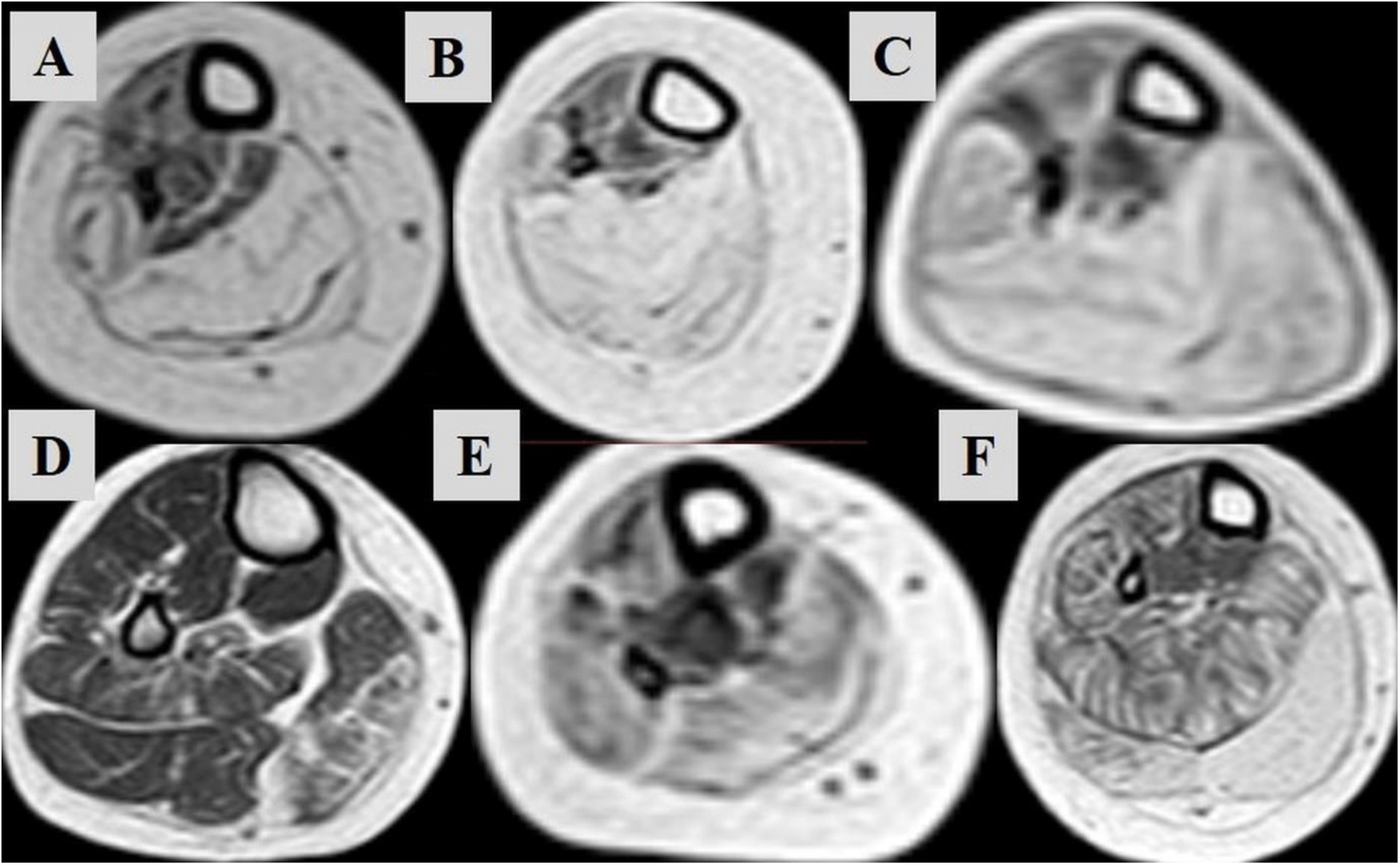
Axial T1WI of the calf in three different patients with LAMA 2 mutation CMD (A, B, C) & three different control (D: collagen 6 deficiency, E: titin deficiency, F: sarcoglyconopathy) showing affection of the soleus & gastrocnemius muscles with relatively spared extensor muscles of the ankle that is less constant in the control cases.

In our study specific patterns were demonstrated within some muscles e.g. deltoid, rectus femoris, sartorius, gracilis, gastrocnemius and erector spinae muscles [Figure 6]. Additionally, the pattern of muscle involvement in the index group differed from that in the control group. There was a statistically significant difference between the two groups in regard to the fatty infiltration of the neck, serratus anterior, flexor & extensor forearm, gluteus maximus, gluteus medius, gastrocnemius and soleus muscles P<0.01. We also found a statistically significant difference between the two groups in regard to the fatty infiltration of the inter-costal, rotator cuff, deltoid and triceps muscles [Table 2]. Additionally, we found a highly statistically significant difference between the fatty infiltration of index and control groups in regard to the neck, shoulder girdle, forearm, pelvic girdle, and calf muscles [Table 3]. An example of white matter demyelination changes in brain MRI is demonstrated in a patient from the index group [Figure 7].

**Tables 3.** Comparison between the index & control groups regarding presence of fatty infiltration in different muscle groups

| Fatty infiltration | Cases<br>N=10 |  | Control<br>N=13 |  | t | P |
| --- | --- | --- | --- | --- | --- | --- |
|  | Mean | SD | Mean | SD |  |  |
| Neck | **2.0** | 1.1 | 0.23 | 0.5 | 4.7 | **0.000\*\*** |
| Abdominal wall | **3.3** | 0.9 | 2.3 | 1.4 | 1.9 | 0.06 |
| Chest wall | **9.9** | 2.1 | 6.0 | 4.3 | 2.5 | **0.01\*** |
| Trunk muscles | 22.9 | 4.8 | 17.3 | 9.7 | 1.7 | 0.09 |
| Shoulder girdle | **11.9** | 3.3 | 7.0 | 4.5 | 2.9 | **0.008\*\*** |
| Upper arm | 6.3 | 2.3 | 5.0 | 2.8 | 0.02 | 0.9 |
| Forearm muscles | **4.7** | 1.4 | 2.0 | 0.7 | 5.9 | **0.000\*\*** |
| Pelvic girdle | **10.8** | 1.5 | 7.2 | 3.3 | 3.4 | **0.003\*\*** |
| Thigh muscle | 12.1 | 2.5 | 9.6 | 5.8 | 1.2 | 0.2 |
| Hamstrings | 8.6 | 1.5 | 7.1 | 2.5 | 1.5 | 0.1 |
| Calf muscles | **11.4** | 1.2 | 7.7 | 3.0 | 3.8 | **0.001\*\*** |
| Leg muscles | 13.0 | 4.3 | 9.7 | 5.4 | 1.5 | 0.1 |

**Figure 6.**
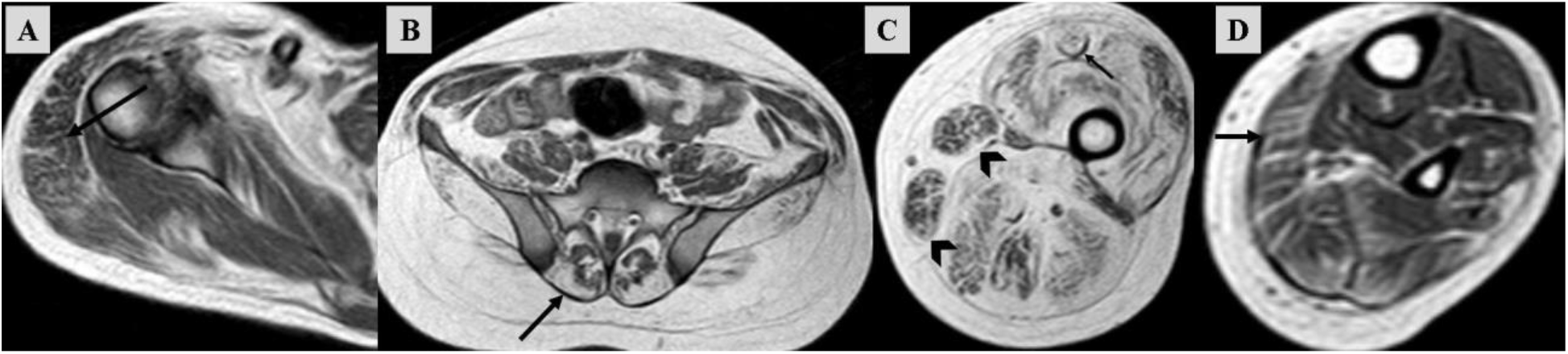
Selected Axial T1WI of the shoulder (A), gluteal (B), thigh (C) and calf (D) regions in patients with LAMA 2 mutation CMD showing granular pattern of affection in the deltoid muscle (arrow in A), sartorius and gracilis muscles (arrow heads in C), more affection of the lateral aspects of the erector spinae muscles (arrow in B), ring like sign within the rectus femoris muscles (small arrows in C) and striated pattern of gastrocnemius muscle affection (arrow in D).

**Figure 7.**
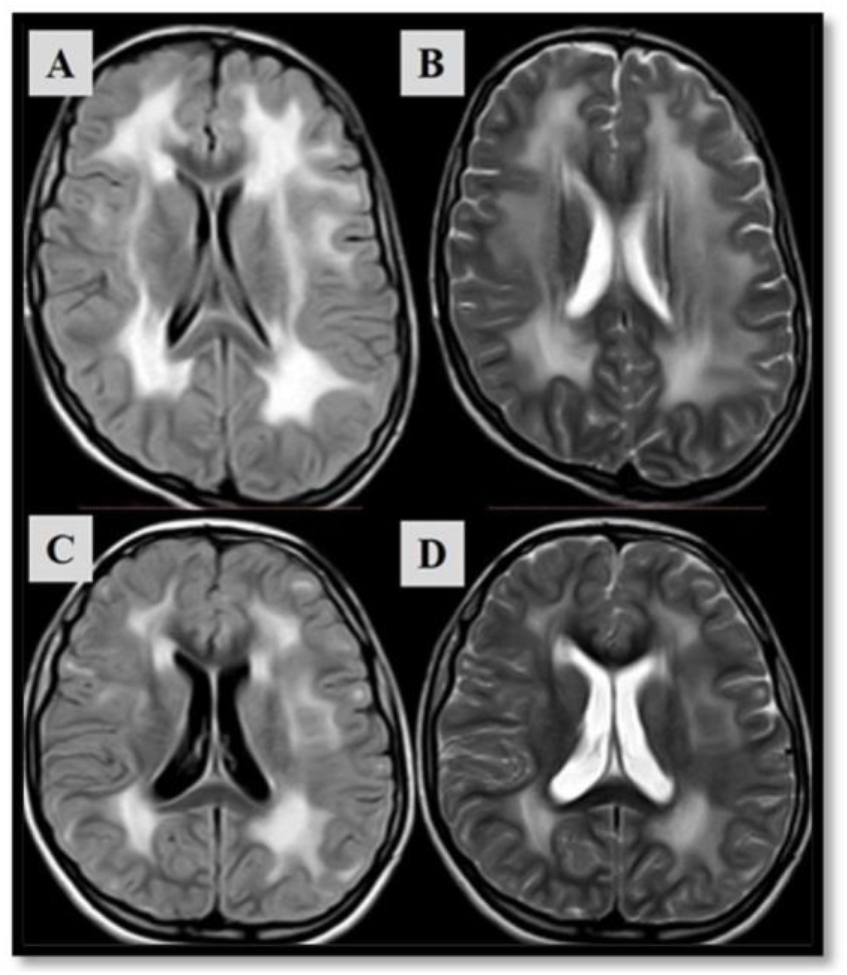
Axial FLAIR (A, C) and T2WI (B, D) of two different LAMA 2 mutation CMD patients showing white matter demyelination change (Leukodystrophy).

## Discussion

Generally, the diagnosis and differentiation of various subtypes of genetic muscle diseases in general and CMD subtypes in specific is a daunting task. Muscle MRI is a promising tool in this regard as it provides high quality anatomical-pathological details and allows for systematic and semi quantitative evaluation of individual muscles of the whole body [3, 13, 19-23]. The overall pattern of whole-body muscle involvement is more helpful than the degree of involvement of an individual muscle. And is capable of producing diagnostically helpful disease-specific and subtype-specific correlations. The pattern of identification of muscle involvement may also be influenced by the characteristic quantitative and qualitative involvement of an individual muscle and chronological order of its involvement. Likewise, the characteristic sparing –relative or absolute- of individual muscle(s) or muscle groups is equally important to the pattern of muscle involvement and an integral part of the muscle MRI signature [19-31]. This may be particularly relevant to early detection of genetic muscle diseases before the full blown muscle MRI signature is apparent in the later stages [24-31].

The above observations have also been a helpful guide to differential diagnosis of CMD subtypes including merosin-deficient CMD. For example, the characteristic pattern of muscle affection in collagen VI-related myopathies shows diffuse affection of the thigh muscles, the quadriceps muscle is the most severely affected muscle while the sartorius, gracilis and adductor longus muscles are relatively spared, a characteristic central signal abnormality is usually seen within the rectus femoris muscle “pseudocollagen sign” or “ring-like pattern” [6, 31]. This sign has been suggested to be correlated to disease severity and level of disability [30]. Additionally, in myotonic muscular dystrophy, among upper limb muscles, the flexor digitorum profundus muscle was most affected, followed by the adductor pollicis longus and the extensor pollicis longus muscles [32, 33].

Although the above patterns share some features with the pattern we noticed among merosindeficient CMD cases, some notable differences remain. For example, the quadriceps muscle, semitendinosus muscle and the extensor muscles of the trunk are not among the commonly and severely affected muscles in our study in contrast to the rotator cuff, calf and peroneal muscles. While some muscle MRI abnormalities were unique to merosin-deficient CMD, others were sensitive yet not specific i.e. shared by other types of CMD as collagen VI-related myopathies. In collagen VI-related myopathy, fatty and connective tissue muscle replacement usually starts around the fascia with this fining more obvious at the rectus femoris and vastus lateralis muscles. Although this “ring-like” pattern of muscle affection is helpful, it is usually absent in advanced cases [6, 34]. In our study specific patterns of involvement were noticed within the muscle itself as “ring-like” affection in rectus femoris muscle, “granular” pattern of affection in deltoid, sartorius and gracilis muscles, “striated” pattern in gastrocnemius muscle and “lateral more than medial” affection in erector spinae muscle [Figure 6].

### Strengths and limitations

The inclusion of a control group with other genetic muscle diseases put more strength to the validity of the results and conclusions. The diversity of the clinical profiles and severities of merosin-deficient CMD patients allowed us to cover a broad scope of pathologic patterns of involvement as per whole-body muscle MRI. Additionally, the inclusion of genetically confirmed cases resolves diagnostic uncertainties. We acknowledge limitations of this study. This was a descriptive retrospective study and therefore we were unable to accurately report on disease progression and select a truly homogenous study sample in regard to patient and disease characteristics. Longitudinal/natural history [3] studies employing whole-body muscle MRI among other modalities have the potential to predict prognosis and signs of disease severity [35, 36], establish relevant functional-imaging correlations and evaluate the responses to therapeutic agents [37-40]. However, the emergence of a fairly characteristic pattern of whole-body muscle MRI changes in our study is an important step to achieve these previous goals. An additional limitation was that some of the whole-body muscle MRI outcome measures were semi-quantitative.

## Conclusions

There is emerging evidence to suggest that whole-body muscle MRI can become a useful contributor to the differential diagnosis of merosin-deficient CMD. The presence of fairly characteristic, sensitive, and specific whole-body muscle MRI findings was documented. The study may be considered as a preliminary step of arriving to establish a reliable MRI classification system for children with merosin-deficient CMD. The whole-body muscle MRI findings should be interpreted with reference to the clinical and molecular context to improve diagnostic accuracy. Larger and longitudinal studies may yet have positive implications for disease tracking and for phenotype-genotype correlations. Future research should focus on establishing correlations between the specific whole-body muscle MRI pattern and the clinical orthopedic manifestations among others.

## Data Availability

The data that support the findings of this study are available from the corresponding author, [HMS] upon reasonable request.

## Acknowledgements

The authors thank Dr Khaled M. Abd Elaziz, Professor of Public Health, Ain Shams University for his help in performing the statistical analysis. This research received no specific grant from any funding agency in the public, commercial or not-for-profit sectors. The authors declare that they have no competing interests. All procedures performed in studies involving human participants were in accordance with the ethical standards of the institutional research committee and with the 1964 Helsinki declaration and its later amendments. The study was approved by the Medical Ethics Research Committee of Faculty of Medicine, Ain Shams University, Egypt number FMASU R 33/2021

## Notes

### Competing Interest Statement

The authors have declared no competing interest.

### Clinical Trial

FMASU R 33/2021

### Clinical Protocols

http://www.aacpdm.org/meetings/2018/program/posters

### Author Declarations

The study was approved by the Medical Ethics Research Committee of Faculty of Medicine, Ain Shams University, Egypt number FMASU R 33/2021

